# A transversal overview of Intensive Care Units environmental microbiome and antimicrobial resistance profile in Brazil

**DOI:** 10.1101/2024.01.29.24301943

**Authors:** Daniela Carolina de Bastiani, Claudia Vallone Silva, Ana Paula Christoff, Giuliano Netto Flores Cruz, Leonardo Daniel Tavares, Luana Silva Rodrigues de Araújo, Bruno Martins Tomazini, Beatriz Arns, Filipe Teixeira Piastrelli, Alexandre Biasi Cavalcanti, Luiz Felipe Valter de Oliveira, Adriano Jose Pereira, the IMPACTO MR investigators

**Affiliations:** BiomeHub Biotechnologies, Florianópolis, SC, Brazil; Big Data Department - Hospital Israelita Albert Einstein, SP, Brazil; Hospital Sírio Libanês, SP, Brazil; Hcor Research Institute, HCor, SP, Brazil; Hospital Moinhos de Vento, RS, Brazil; Hospital Alemão Oswaldo Cruz, SP, Brazil

**Author notes:** These authors contributed equally to this work and share first authorship (Daniela Carolina de Bastiani, Claudia Vallone Silva, Ana Paula Christoff). **Correspondence:** Adriano José Pereira.

**Keywords:** microbiome, bacteria, antimicrobial resistance (AMR) gene, NGS – next generation sequencing, hospital surfaces, 16S rRNA amplicon

## Abstract

**Introduction:** Infections acquired during healthcare setting stay pose significant public health threats. These infections are known as Healthcare-Associated Infections (HAI), mostly caused by pathogenic bacteria, which exhibit a wide range of antimicrobial resistance.

**Objective:** Characterize the microbiome and antimicrobial resistance genes present in high-touch Intensive Care Unit (ICU) surfaces, and to identify the potential contamination of the sanitizers/processes used to clean hospital surfaces.

**Methods:** In this national, multicenter, observational, and prospective cohort, bacterial profiles and antimicrobial resistance genes from 41 hospitals across 16 Brazilian states were evaluated. Using high-throughput 16S rRNA amplicon sequencing and real-time PCR, the bacterial abundance and resistance genes presence were analyzed in both ICU environments and cleaning products.

**Results:** We identified a wide diversity of microbial populations with a recurring presence of HAI-related bacteria among most of the hospitals. The median bacterial positivity rate in surface samples was high (88.24%), varying from 21.62% to 100% in different hospitals. Hospitals with the highest bacterial load in samples were also the ones with highest HAI-related abundances. *Streptococcus spp*, *Corynebacterium spp*, *Staphylococcus spp*, *Bacillus spp*, *Acinetobacter spp,* and bacteria from the Flavobacteriaceae family were the microorganisms most found across all hospitals. Despite each hospital particularities in bacterial composition, clustering profiles were found for surfaces and locations in the ICU. Antimicrobial resistance genes *mecA*, *bla*_KPC-like_, *bla*_NDM-like_, and *bla*_OXA-23-like_ were the most frequently detected in surface samples. A wide variety of sanitizers were collected, with 19 different active principles in-use, and 21% of the solutions collected showed viable bacterial growth with antimicrobial resistance genes detected.

**Conclusion:** This study demonstrated a diverse and spread pattern of bacteria and antimicrobial resistance genes covering a large part of the national territory in ICU surface samples and in sanitizers solutions. This data should contribute to the adoption of surveillance programs to improve HAI control strategies and demonstrate that large-scale epidemiology studies must be performed to further understand the implications of bacterial contamination in hospital surfaces and sanitizer solutions.

## Introduction

Healthcare-associated infections (HAI) are infections acquired during hospital or another healthcare setting stay, and pose significant public health threats, particularly in low- or middle-income countries (LMIC) like Brazil (1,2). According to a report published by the Joint Commission, HAI rates are close to 7.1% in hospitalized patients in Europe, 4.5% in the US and 15.5% in LMIC countries, and when it comes to ICU infections, LMIC have rates more than 3 times higher than in the US (13.6/1000 patient-days versus 47.9/1000 patient-days) (3).

Pathogenic bacteria exhibit a wide range of antimicrobial resistance (AMR) and have the potential of carrying multidrug-resistance (MDR) genes (4,5). These aggravating factors increase the HAI burden with serious implications for patient health leading to increased length of hospital stay and mortality, also driving up healthcare expenses (6). Therefore, close monitoring and improvement of surveillance programs are necessary.

Over the past years, it has been shown that healthcare workers behavior, patients characteristics, and factors related to the hospital environments, including surfaces, play a critical role in the dissemination of hospital pathogens (7–9). Studies have shown the existence and enduring presence of specific bacterial pathogens, such as *Pseudomonas spp*, *Acinetobacter spp*, *Staphylococcus spp*, *Corynebacterium spp*, *Sphingomonas spp*, and *Clostridium spp* on hospital surfaces (10–13). Furthermore, when a particular patient is exposed to an environment previously occupied by a MDR colonized patient, this new patient is susceptible to colonization by the same organisms, suggesting that the cleanliness of the healthcare environment seems to be an important factor preventing MDR bacteria transmission (14,15). However, more robust evidence is still lacking whether more aggressive strategies to disinfect Intensive Care Units (ICU) and other hospital environments could reduce rates of HAIs.

Hospital microbiome studies using culture independent methods, such as high-throughput sequencing (HTS), conducted in healthcare institutions (16–18), enable a large-scale screening of microorganisms directly from collected samples, including those that may not thrive under conventional microbiology conditions (19–23) and contribute to the understanding of crucial aspects related to HAI. Also, these studies provide a more comprehensive view of the microbial profile in the environment, and how the adoption of surveillance programs based on surface DNA HTS can improve effective HAI control strategies (16,23,24).

This study aims to characterize the microbiome of high-touch ICU surfaces, and to map the potential contamination of the sanitizers used to clean hospital surfaces in an upper middle-income country.

## Methods

### Study design

This is a national, multicenter, observational, and prospective cohort conducted in 41 hospitals from 16 different Brazilian states. The project is part of a major initiative called IMPACTO MR program, a nationwide registry and platform for observational studies and trials on HAIs, especially those caused by multidrug-resistant Organisms (MDROs) (25). The study was approved by the Hospital Israelita Albert Einstein (HIAE) - São Paulo - Brazil Ethics Committee (approval number 4.122.595), and also by the Hospital do Coração (HCor) - São Paulo - Brazil Ethics committee (approval number 4.040.974). We invited all 50 ICUs of the IMPACTO MR platform to participate in this data collection, and 38 ICUs accepted the invitation. Also, three ICUs in the State of São Paulo were used to run the pilot study.

### Swab sample collection and DNA extraction

All hospitals were visited by a trained healthcare professional in sample collection, following a standard approach defined by a nurse specialist in Infection Control (CVS) and a specialist Microbiology/Bioinformatics (APC), from October-2020 to January-2021. ICU rooms, nursing stations and prescription areas in the ICU were sampled. For each hospital, 38 swabs (hospital environment samples) from high-touch surfaces, such as medical and hospital equipment, furniture, critical structure points and bed accessories, from ICU common areas, and 5 beds (being 3 during patient care, and 2 after discharge and terminal cleaning) were collected, as described in S1 Table.

Samples were collected using a dry sterile hydraflock swab (Puritan, USA). Prior to sample collection, the swab was moistened with a sterile saline solution (0.9% NaCl). After sample collection, the swab tip was broken down into a microtube containing 800 μL of stabilization solution–ZSample (BiomeHub, BR) (26) that allowed storage and transport up to 30 days at room temperature. The swabs were sent to the laboratory facilities (BiomeHub, BR) to be processed as previously described (26). Briefly, the DNA from the samples was obtained through a thermal lysis (96°C – 10 min) followed by a purification step with magnetic beads (Sera-Mag^TM^ SpeedBeads Carboxylate-Modified Particles, Cytiva, UK). Negative controls (only reagents) were included in each lysis and DNA extraction batch.

### Sanitizer sample collection, bacterial culture, and DNA extraction

Two samples of different sanitizing solutions being used in the hospital routine were collected: one sanitizing solution, the most used in the daily routine cleaning (concurrent) by the nursing team, and another, most used in terminal room cleaning (patient discharge) by the hygiene team. Samples were collected in 200 mL sterile bottles, directly from the sanitizer in-use container from each hospital. The intention was to be representative of the last stage before the sanitizer reaches the targeted surface (contamination in the process of use). They were transported at room temperature, as indicated in the storage instructions for the original sanitizer product and forwarded to an ISO/IEC 17025 accredited microbiology laboratory to perform growth and total count analysis of mesophilic aerobic bacteria. Sanitizing solutions were sent for microbiological culture, given the need to neutralize chemical compounds present in sanitizers that could interfere with a direct DNA extraction approach.

Giving the sanitizers chemical diversity, the laboratory inoculated a positive control sample (bacteria positive) along with each sanitizer culture sample. This allowed the confirmation of correct sanitizer active principle inactivation for proper microbial growth. Otherwise, culture results were reported as inconclusive, due to the lack of sanitizer neutralization and possible interference in the results (false negative). When a culture sample turned to be positive (with microbial growth), the pool or isolated microorganisms that grew for each sanitizer were sent back to the laboratory facilities (BiomeHub, BR) to be identified with high-throughput amplicon 16S rRNA sequencing, and also resistance genes Real-Time PCR (RGene - BiomeHub, SC) analysis. The DNA extraction for culture isolates was carried out as described above.

### Library preparation and DNA sequencing

The 16S rRNA amplicon sequencing libraries were prepared using the V3/V4 primers (341F CCTACGGGRSGCAGCAG and 806R GGACTACHVGGGTWTCTAAT) (27,28) in a two-step PCR protocol. The first PCR was performed with V3/V4 universal primers containing a partial Illumina adaptor, based on TruSeq structure adapter (Illumina, USA) that allows a second PCR with the indexing sequences similar to procedures described previously (27). Here, combinatorial dual-indexes were added per sample in the second PCR, also performing index switches between runs to avoid cross contaminations. Two microliters of individual sample DNA were used as input in the first PCR reaction. The PCR reactions were carried out using Platinum Taq (Invitrogen, USA). The conditions for PCR1 were: 95°C for 5 min, 25 cycles of 95°C for 45 s, 55°C for 30 s, and 72°C for 45 s and a final extension of 72°C for 2 min for PCR 1. For PCR 2, two microliters of the first PCR were used and the amplification conditions were 95°C for 5 min, 10 cycles of 95°C for 45.s, 66°C for 30.s, and 72°C for 45.s with a final extension of 72°C for 2 min. All PCR reactions were performed in triplicates. The second PCR reactions were cleaned up with magnetic beads (Sera-Mag^TM^ SpeedBeads Carboxylate-Modified Particles, Cytiva, UK). and an equivalent volume of each sample (10 - 30 uL) was added in the sequencing library pool. At each batch of PCR reactions, a negative (blank) control was included (only reagents). The final DNA concentration of the library pool was estimated with Quant-iT Picogreen dsDNA assays (Invitrogen, USA), and then diluted for accurate qPCR quantification using Collibri™ Library Quantification Kit (Invitrogen, USA). The sequencing pool was adjusted to a final concentration of 12 pM (for V2 kits) or 18 pM (for V3 kits) and sequenced in a MiSeq system (Illumina, USA), using the standard Illumina primers provided by the manufacturer kit. Single-end 300 cycle runs were performed using V2×300, V2×300 Micro, or V3×600 sequencing kits (Illumina, USA) with an average sample depth expected of 30k reads per sample.

### DNA sequencing data analysis

The read sequences were analyzed using a bioinformatics pipeline previously described (16,17,26) (BiomeHub, Brazil-hospital_microbiome_rrna16s: v1). Illumina FASTQ files had the primers trimmed and their accumulated error was assessed (26). Reads were analyzed with the Deblur package (29) to discard potentially erroneous reads and then reads with identical sequences were grouped into oligotypes (clusters with 100% identity, ASVs amplicon sequencing variants). Next, VSEARCH (30) was used to remove chimeric amplicons. An additional filter was implemented to remove oligotypes below the frequency cutoff of 0.2% in the final sample counts. We also implemented a negative control filter, since hospital microbiomes generally are low biomass samples (26). For each processing batch, negative controls (reagent blanks) were included during both DNA extraction and PCR reactions. If any oligotype was recovered in the negative control results, they were checked against the samples and automatically discarded from the results if their abundance (in number of reads) was no greater than two times their respective counts in the controls. The remaining oligotypes in the samples were used for taxonomic assignment with the BLAST tool (31) against a reference genomic database (encoderef16s_rev6_190325). This reference database comprised complete and draft bacterial genomes, with an emphasis on clinically relevant bacteria, obtained from NCBI. It is composed of 11,750 sequences including 1,843 unique different bacterial taxonomies.

Taxonomy was assigned to each oligotype (ASV) using a lowest common ancestor (LCA) algorithm. If more than one reference can be assigned to the same oligotype with equivalent similarity and coverage metrics, the taxonomic assignment algorithm leads the taxonomy to the lowest level of possible unambiguous resolution (genus, family, order, class, phylum or kingdom), according to the similarity thresholds (32).

After a quality check of the final yield, the resulting oligotype tables were processed as previously described (26). Oligotype sequences served as input for FastTree 2.1 software (33) to construct phylogenetic trees. Subsequent analyses were performed using R (version 3.6.0) and the Phyloseq package (34). Alpha diversity analysis included the Shannon diversity index and observed richness. Beta diversity employed Principal Coordinate Analysis with Bray-Curtis dissimilarity computed from proportion-normalized data.

### RGene–antimicrobial resistance gene analysis

A panel including relevant β-lactamases, Vancomycin and Methicillin antimicrobial resistance genes was assembled. Tested genes were: *bla*_CTX-M-1_ group, *bla*_CTX-M-2_ group, *bla*_CTX-M-8_ group, *bla*_CTX-M-9_ group, *bla*_KPC-like_, *bla*_NDM-like_, *bla*_SPM-like_, *bla*_OXA-23-like_, *vanA*, *vanB* and *mecA*. The detection was performed using Real-Time PCR with QSY hydrolysis probes labeled with FAM®, VIC® and NED® (Applied Biosystems, USA). To test primer and probe efficiency we used bacterial strains containing the resistance genes of interest (kindly provided by Prof. Dr. Ana Cristina Gales). The aforementioned bacterial strains were also included in each PCR run as positive controls. Real-Time PCR reactions were conducted using 10 μL of final volume per sample, containing 2 μL of the same previously sequenced DNA samples, 0.2 U Platinum Taq, 1 X Buffer, 3 mM MgCl2, 0.1 mM dNTP, 0.12 X ROX and 0.2 μM of each forward and reverse specific primer following the thermal conditions: 95°C for 5 min with 35 cycles of 95°C for 15s, 60°C for 30s and 72°C for 30s. Negative (reagent blanks) reaction controls were included in all the assays. All the samples were analyzed in experimental triplicates. Real-Time reactions were performed in QuantStudio 6 Pro and QuantStudio 5 384 Real-Time PCR Systems (Applied Biosystems, USA). Samples were considered positive when at least two of the experimental replicates were below the quantification cycle 33 using an experimental threshold of 0.05.

## Results

### Environmental samples and high-throughput amplicon sequencing

In total, 1492 hospital surface samples were collected from the 41 studied ICUs (38 ICUs from the IMPACTO-MR program and 3 ICUs from the pilot study). Not all 38 expected samples could be collected in all hospitals, with each hospital providing between 25 and 38 surface samples, along with 78 in-use sanitizer samples, 2 from each hospital (2 samples were not collected and 2 leaked during transport).

Both total microbial load and sample positivity proportions varied greatly across hospitals. **Fig. 1A** shows the log10-transformed total sequence reads from each hospital. The bacterial positivity rate had median values of 88.24% and varies from 21.62% (H1) to 100% (H17, H25, H28, H42) in samples from each hospital, seemingly unrelated to total microbial load. We observed the same pattern when considering 17 bacteria from a restricted group of interest in healthcare-associated infections, including *Acinetobacter baumannii*, *Burkholderia cenocepacia*, *Burkholderia cepacia*, *Clostridioides difficile*, *Corynebacterium spp*, *Enterobacteriaceae*, *Enterococcus faecalis*, *Enterococcus faecium*, *Escherichia coli*, *Klebsiella pneumoniae*, *Proteus mirabilis*, *Pseudomonas aeruginosa*, *Pseudomonas putida*, *Staphylococcus aureus*, *Staphylococcus epidermidis*, *Staphylococcus hominis,* and *Stenotrophomonas maltophilia* (**Fig. 1B**). The top 5 hospitals for total bacterial load higher than 10^4^ reads are the same as the top 5 considering only the group of 17 specific HAI-related bacteria (H17, H22, H30, H36 and H42). Also, the hospitals with lower amounts of HAI-related bacteria are among the hospitals with the lower medians for total bacterial load (H24, H31 and H38).

**Figure 1.**
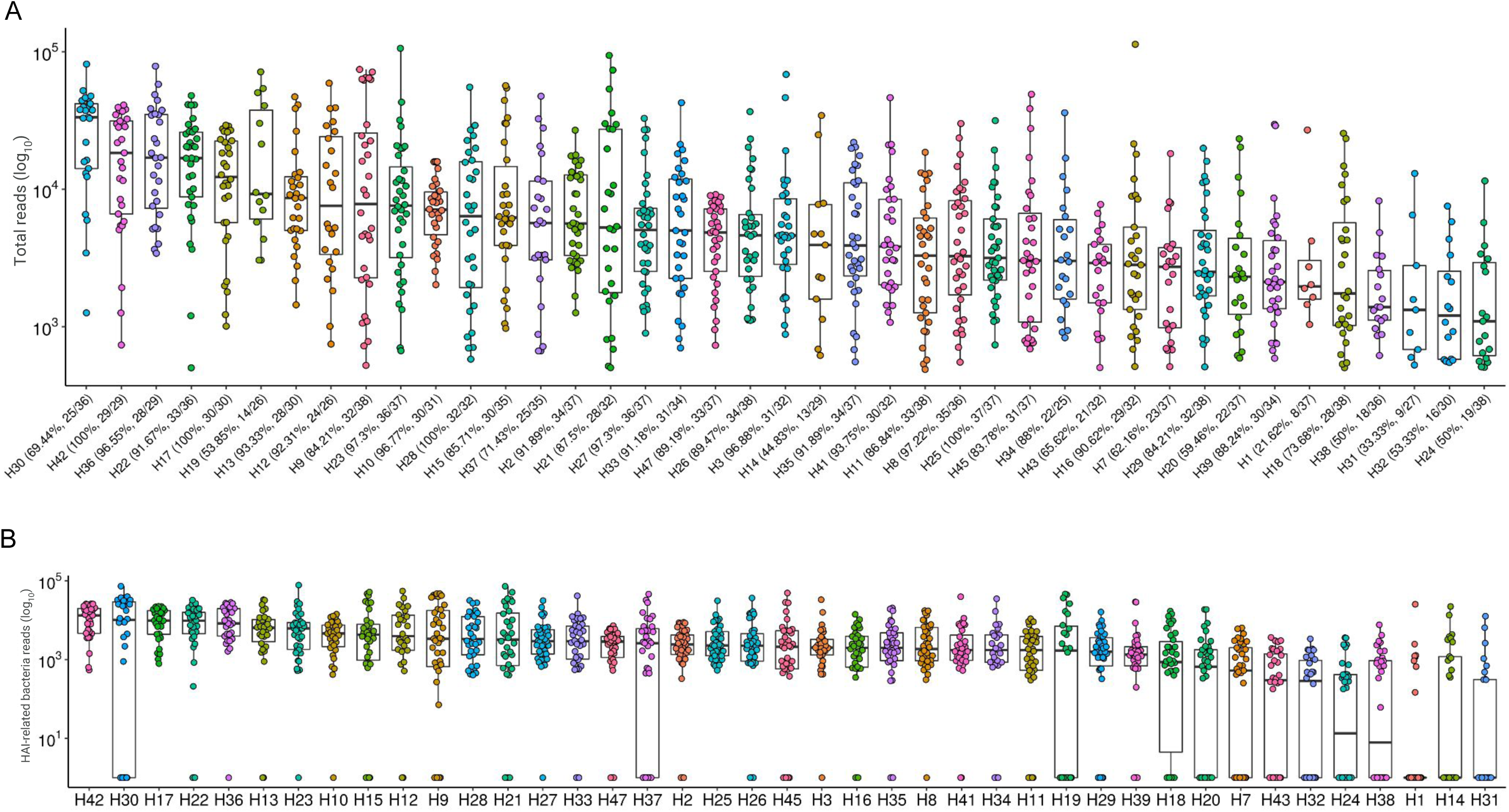
Total sequenced reads in environment samples along 41 hospitals. (A) Total sequenced reads (library size) in log_10_ scale are represented by boxplots with the median bacteria reads for each hospital environment samples collected. Bacterial positivity rate in samples from each hospital is represented by percentage values in the x axis. (B) Total sequence reads for each sample, considering only HAI-related bacteria by hospital.

### Bacterial profiling from the hospital surfaces

This study employed high-throughput amplicon sequencing to identify bacterial taxonomies, which revealed a rich diversity of microbial populations. In order to understand the dispersion of bacteria within the hospitals, the average abundances of the oligotypes were used to plot a heat map. The analysis showed the widespread presence of taxa such as *Streptococcus spp*, *Corynebacterium spp*, *S. epidermidis*, *Flavobacteriaceae*, *Bacillus spp*, and *A. baumannii* across all 41 hospitals (**Fig. 2**). Furthermore, the investigation into the bacterial composition within-hospital environment identified four major distinct clusters characterized by similar positivity patterns of microbial taxa. This hierarchical clustering pattern indicated the existence of common bacterial profiles (presence/absence) associated with different hospitals, as in cluster 1, hospitals with samples highly positive for *Streptococcus spp*, *Corynebacterium spp and S. epidermidis*, or cluster 2 including hospitals with high positivity rates for *Streptococcus spp, Flavobacteriaceae, Bacillus spp, Xanthomonadaceae, Bacillaceae* and *Bordetella spp*. Cluster 3 includes the hospitals with the lowest bacterial positivity rates, and cluster 4 is represented by hospitals with high positivity rates for *Streptococcus*, *Corynebacterium*, *S. epidermidis*, *A. baumannii*, *Pseudomonas*, *Acinetobacter*, *Staphylococcus* and *S. haemolyticus*.

**Figure 2.**
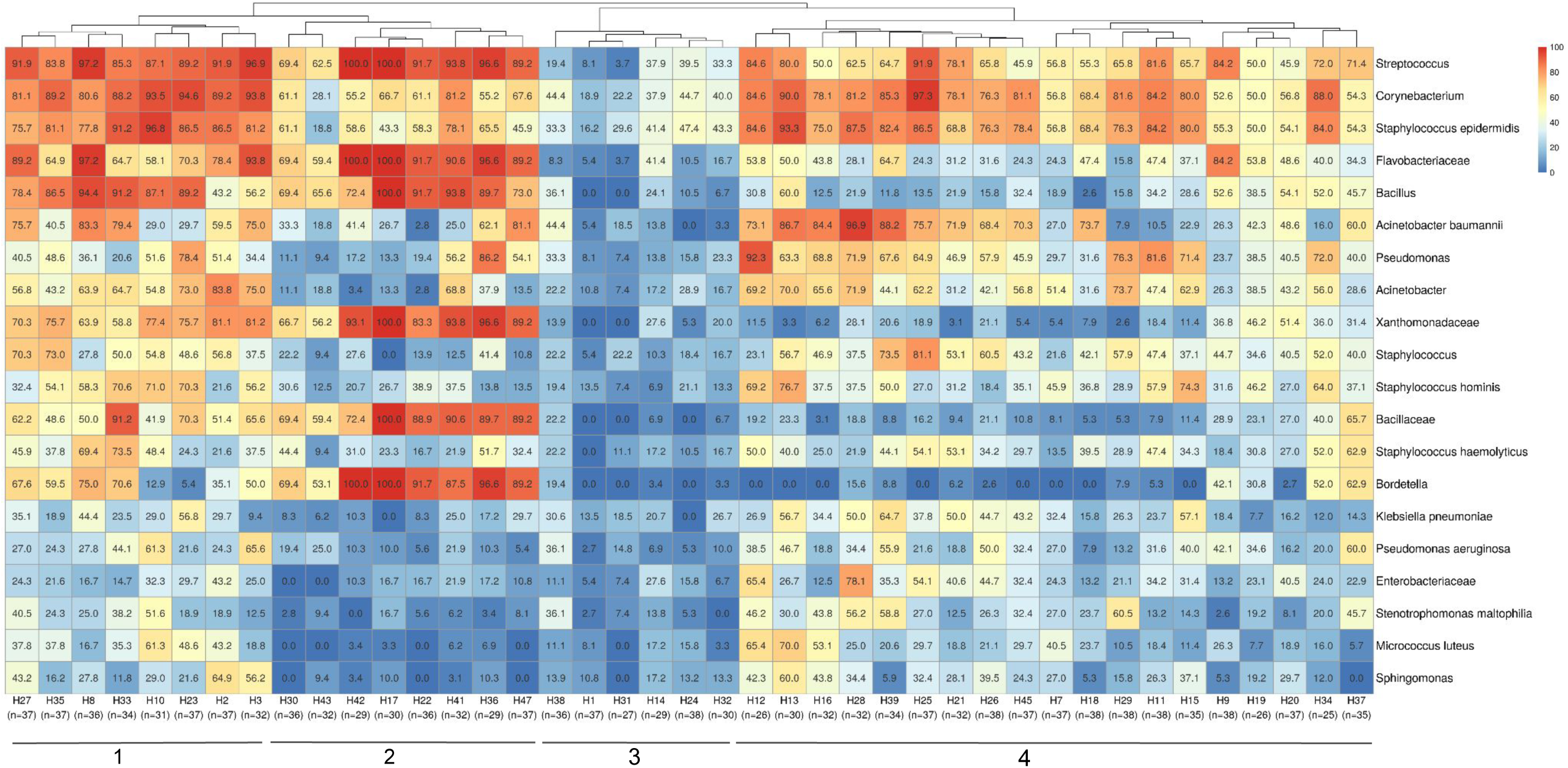
Hospital clustering profiles based on bacterial positivity rates. Most abundant bacteria detected in samples are demonstrated by the heatmap color scales and percentages values representing the proportion of positivity (from 100%, in red, to 0%, in dark blue) in samples from each hospital. The number of analyzed samples for each hospital is indicated below their identification. Also, the four major clustering groups are highlighted by numbers 1-4 at the bottom.

Considering the previously selected 17 specific HAI-related bacteria, we assessed the prevalence of positive samples for these specific taxa in each hospital (**Fig. 3**). Hierarchical clustering showed the segregation of two main hospital groups, based on sample positivity rates. Predominantly, cluster 1 presented lower proportions of positive samples for *S. epidermidis*, *Corynebacterium app*, *A. baumanii, S. hominis, K. pneumoniae and P. aeruginosa* than hospitals in cluster 2. The most prevalent HAI-related bacteria detected in hospitals were *Corynebacterium spp* and *S. epidermidis*.

**Figure 3.**
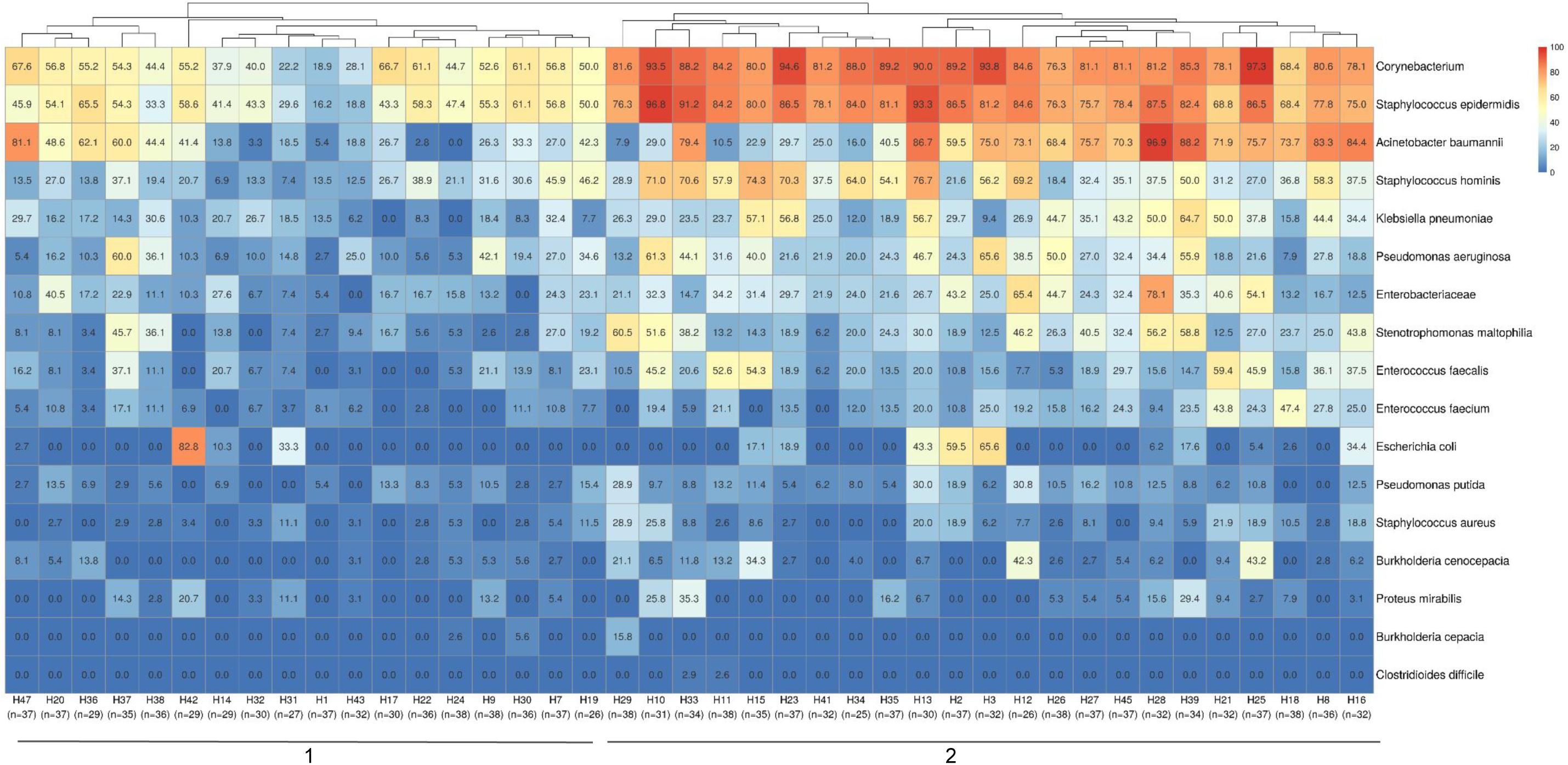
Hospital clustering by HAI relevant bacteria positivity rates in samples. The heatmap shows a specific group of 17 important HAI-related bacteria (*Acinetobacter baumannii*, *Burkholderia cenocepacia*, *Burkholderia cepacia*, *Clostridioides difficile*, *Corynebacterium*, *Enterobacteriaceae*, *Enterococcus faecalis*, *Enterococcus faecium*, *Escherichia coli*, *Klebsiella pneumoniae*, *Proteus mirabilis*, *Pseudomonas aeruginosa*, *Pseudomonas putida*, *Staphylococcus aureus*, *Staphylococcus epidermidis*, *Staphylococcus hominis* and *Stenotrophomonas maltophilia*) selected to observe their distribution among the hospitals in the study. Two major clustering groups could be observed and are indicated at the bottom of the figure (1–2), along with the identification of each hospital and the number of samples analyzed in each one. Samples positivity rates for each bacteria identified in each hospital can be observed by positivity rate values inside the boxes and color scales (0 to 100%).

Examining the surfaces in each hospital environment revealed a diverse bacterial profile and prevalence, in samples for total bacteria detected (**Supp. fig. 1**), and also different positivity rates among hospitals for the specific group (filter) of selected HAI-related bacteria (**Supp. fig. 2**). Considering the surfaces of all hospitals in one analysis for the HAI-related bacteria group (**Fig. 4A**), a hierarchical clustering highlighted more similarities between bacterial prevalences in the hygiene material (mop handle with squeegee and cleaning cart) and the nursing station faucet plus soap dispenser sites. Also, some specific bed sites - gas ruler plus flowmeter, monitor buttons plus infusion pump, and bed rails - were more similar in bacterial composition and prevalence. Nursing station counters were more related to medical prescription sites as well as to meal and/or procedures tables (that were mainly stored in nursing stations). However, these nursing and prescription sites were also more similar to the remaining nursing (medication area and alcohol dispenser) and prescription (computer keyboard and mouse) sites, as well as with bed IV stands, curtains, partitions and door knobs. Despite this general profile characterization, each hospital has its particular bacterial dispersion, ranging from almost all negative samples, except in beds occupied by patients (beds 1, 2 and 3, **Fig. 4B**), to highly dispersed bacteria across samples (**Fig. 4C**). More specific dispersion profiles were also observed, such as for *C. difficile,* found only in bed 1 from H33 hospital (**Fig. 4C**) and bed 3 from H11 (**Fig. 4D**), or *S. aureus* only on medical prescription surfaces from H11.

**Figure 4.**
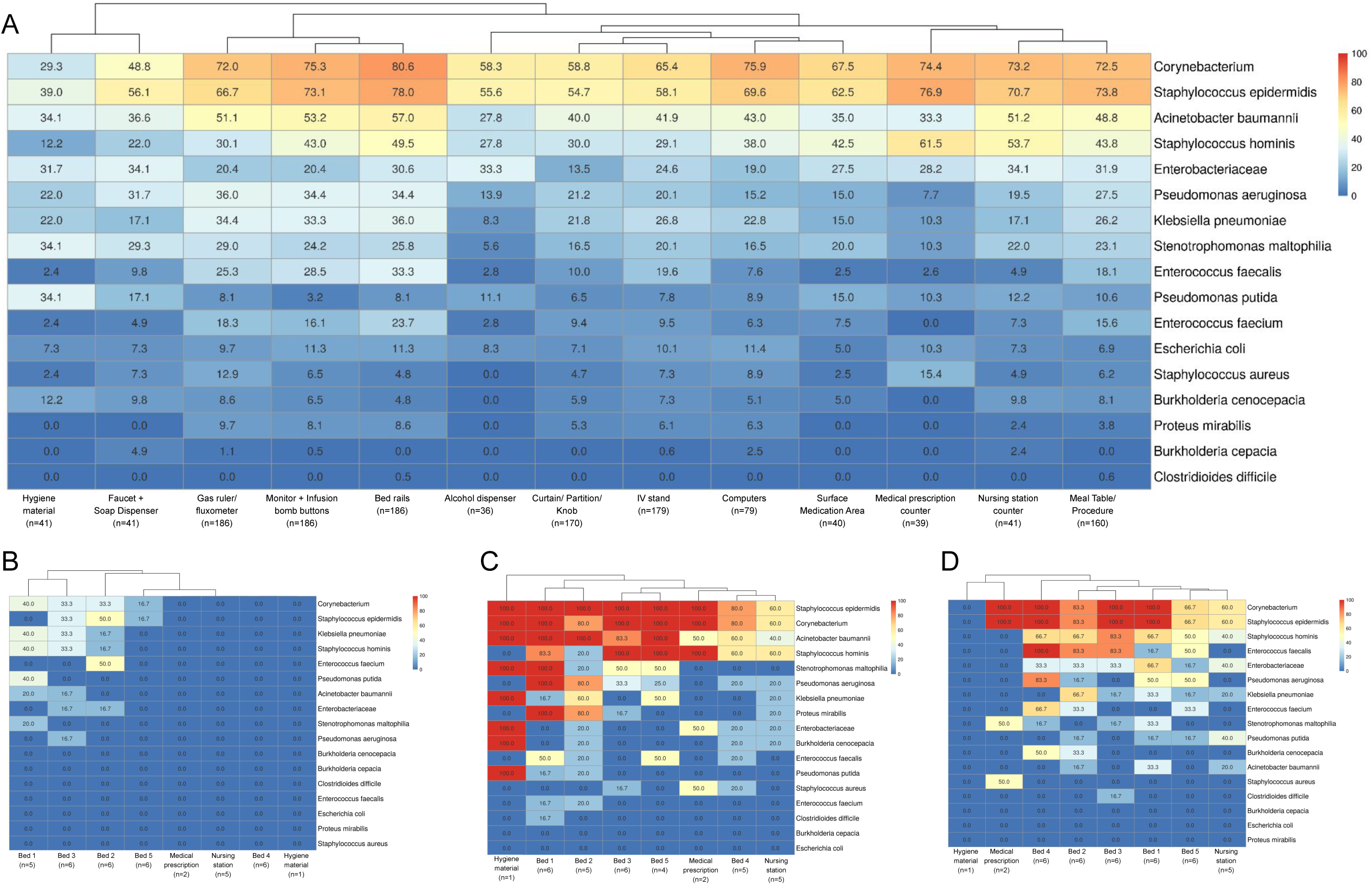
Bacterial positivity rates in surface samples. **(A)** Considering the 17 HAI-related bacteria (described previously), a heatmap of their positivity rates in hospital surface types was shown. The number of samples included for each type of surface is indicated below their description in the figure. Figures B, C and D show the differential profiles for bacterial positivity rates in the hospitals included in this study. The heatmaps indicated the bacterial positivity rates inside boxes and a color scale from 0 to 100% for sample locations. (B) A hospital with low bacterial contamination and dispersion patterns (hospital H1), only concentrated in beds 1, 2 and 3, which are the ones occupied by patients at the time of sample collection. (C) A high bacterial contamination and dispersion among almost all samples (hospital H33) and (D) hospitals such as H11 with specific bacterial dispersion profiles as for *S. aureus* detected only in one medical prescription site (50% from 2 analyzed samples) or *C. difficile* found only in bed 3 samples (16.3% from 6 analyzed samples).

Diversity metrics, Shannon and Richness indexes were computed per hospital, considering all samples and presented maximum values of 4.66 and 145 respectively (**Supp. fig. 3A and 3B**). Beta-diversity using Principal Coordinate Analysis (PCoA) with Bray-Curtis dissimilarity was performed to identify possible similarities (grouping) among hospitals (**Supp. fig. 4**) or different country states (**Supp. fig. 5**). No particularities from bacterial profiles in different hospitals or country regions were found significant.

### Bacterial profiling from in-use hospital sanitizers

A total of 78 in-use sanitizer samples (diluted), two from each hospital, were analyzed, one being used in the concurrent cleaning by the nursing team, and another in terminal cleaning by the hygiene team. From these, there were 42 different commercially available products and 19 different active principles (**Supp. table 2**) used in different combinations by the two teams in each hospital. The most used sanitizer product by the nursing team was alcohol 70%, while by the hygiene team was the 5th generation quaternary ammonium and biguanide (**Fig. 5**). There is no standardization of hospital cleaning products used in Brazil at the country level, but any product used must be registered with the competent official body (ANVISA) and have its efficacy tested against some reference microorganisms. Each hospital has its own sanitizer (or sanitizer combination) choice. Indeed, among the 41 investigated hospitals, there were only 3 using the same active principle sanitizer combination by both teams (H26, H33, H37).

**Figure 5.**
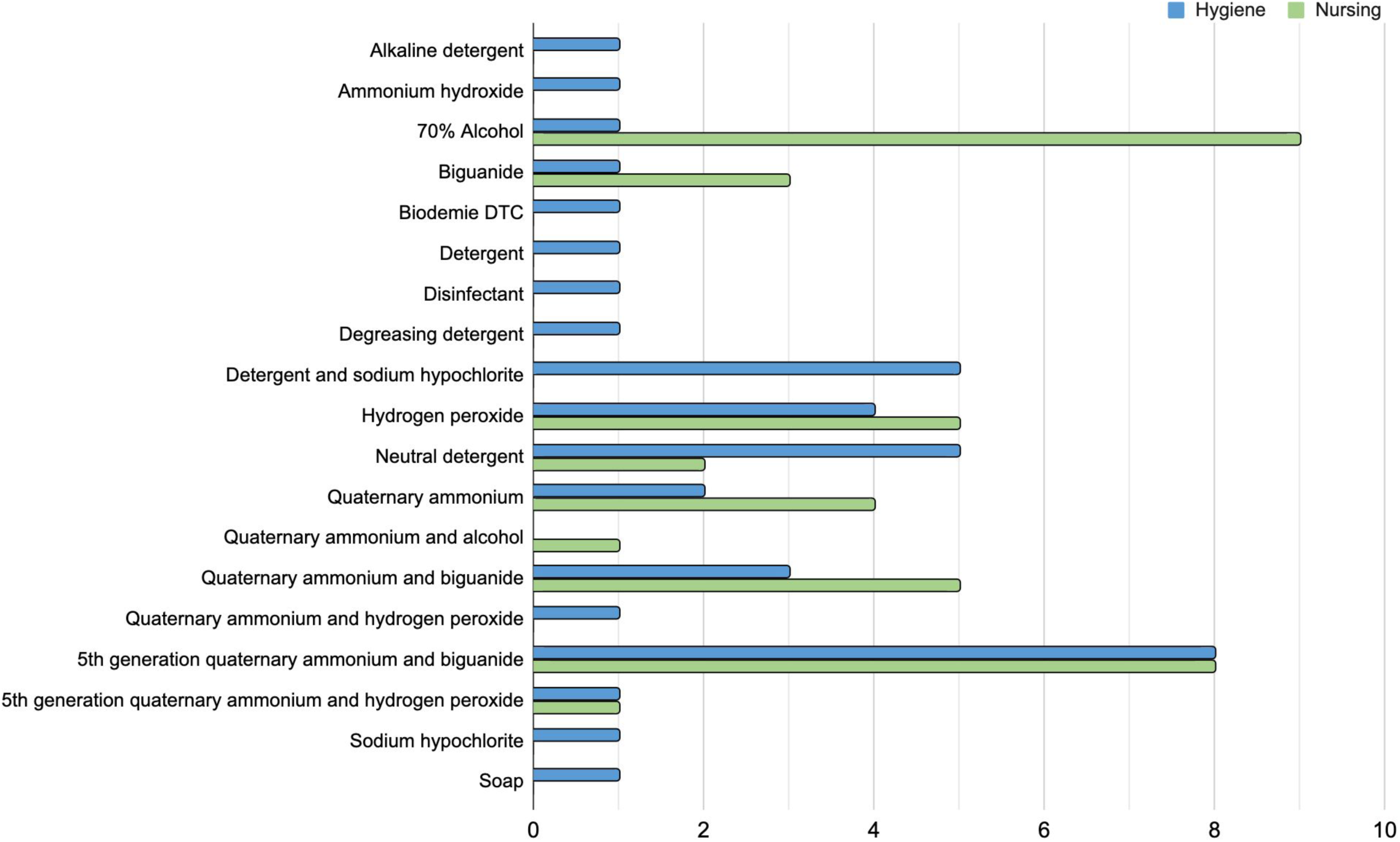
Sanitizers in use by Brazilian hospitals. The bar plot indicates all the active principles of in-use sanitizers by the 41 hospitals included in this study. The number of hospitals using a sanitizer solution with the related active principle is separated by the cleaning team who uses it, in blue the hygiene team (terminal cleaning after patient discharge) and in green the nursing team (daily concurrent cleaning while patient is still in bed).

From the 78 in-use sanitizer samples, 17 had mesophilic aerobic bacteria growth, including the following members of HAI-related bacteria: *A. baumannii*, *B. cepacia* complex, *E. coli*, *P. putida* and *S. maltophilia* (<u>Table S3</u>). These bacterial growths varied from 10 CFU/g to >5.0 x10^3^ CFU/g. 51 sanitizer samples had bacterial growth undetected (< 10 CFU/g) and 10 sanitizer samples had inconclusive results due to the failure of active principle inactivation, compromising microbial culture reliability for a true negative result.

### Antimicrobial resistance genes

Antimicrobial resistance genes were investigated in all hospital surface samples and culture-positive sanitizers samples. From all 1492 environmental tested samples, 74.80% were positive for at least one AMR gene. **Figure 6A** shows the prevalence of AMR genes among different hospital environmental collection sites, considering all the hospitals included in the project. The most frequently detected AMR genes were *mecA*, *bla*_KPC-like_, *bla*_NDM-like_ and *bla*_OXA-23-like_, while *bla*_CTX-M-9_ group and *vanB* were not identified in any collected sample. Furthermore, the *mecA* gene was detected in most of the analyzed samples. Terminal cleaning process at patient discharge seems to slightly decrease the prevalence of AMR genes in bed samples, compared to the daily cleaning processes while patients are still in the rooms (**Fig. 6B**). From 17 culture-positive sanitizer samples, 3 had AMR genes detected: *bla*_CTX-M-8_ group, *bla*_CTX-M-2_ group and *bla*_NDM-like_ in a neutral detergent from H7, *bla*_CTX-M-8_ group and *bla*_KPC-like_ in an alkaline detergent from H19 and also in a local supplier sanitizer product from H47 (<u>Table S3</u>). These three hospitals from which sanitizers presented resistance genes in the sanitizer solutions also had the same AMR genes detected in environmental surface samples collected by swab, in variable degrees. Hospital H7 had *bla*_NDM-like_ detected in 6 samples (16.2%): in the faucet and soap dispenser, the meal/procedure table, in bed rails, curtain/division or door knobs and monitor and infusion bomb buttons. H19 hospital had *bla*_CTX-M-8_ group AMR gene detected in bed rails and *bla*_KPC-like_ in 16 samples (61.5%), including mostly bed related samples, but also nursing station samples as in the medication area, computer and hygiene material. In H47, *bla*_CTX-M-8_ group was detected in one IV stand and in the gas ruler from two different rooms, and *bla*_KPC-like_ AMR gene was present in 70.3 % of the collected samples in that hospital.

**Figure 6.**
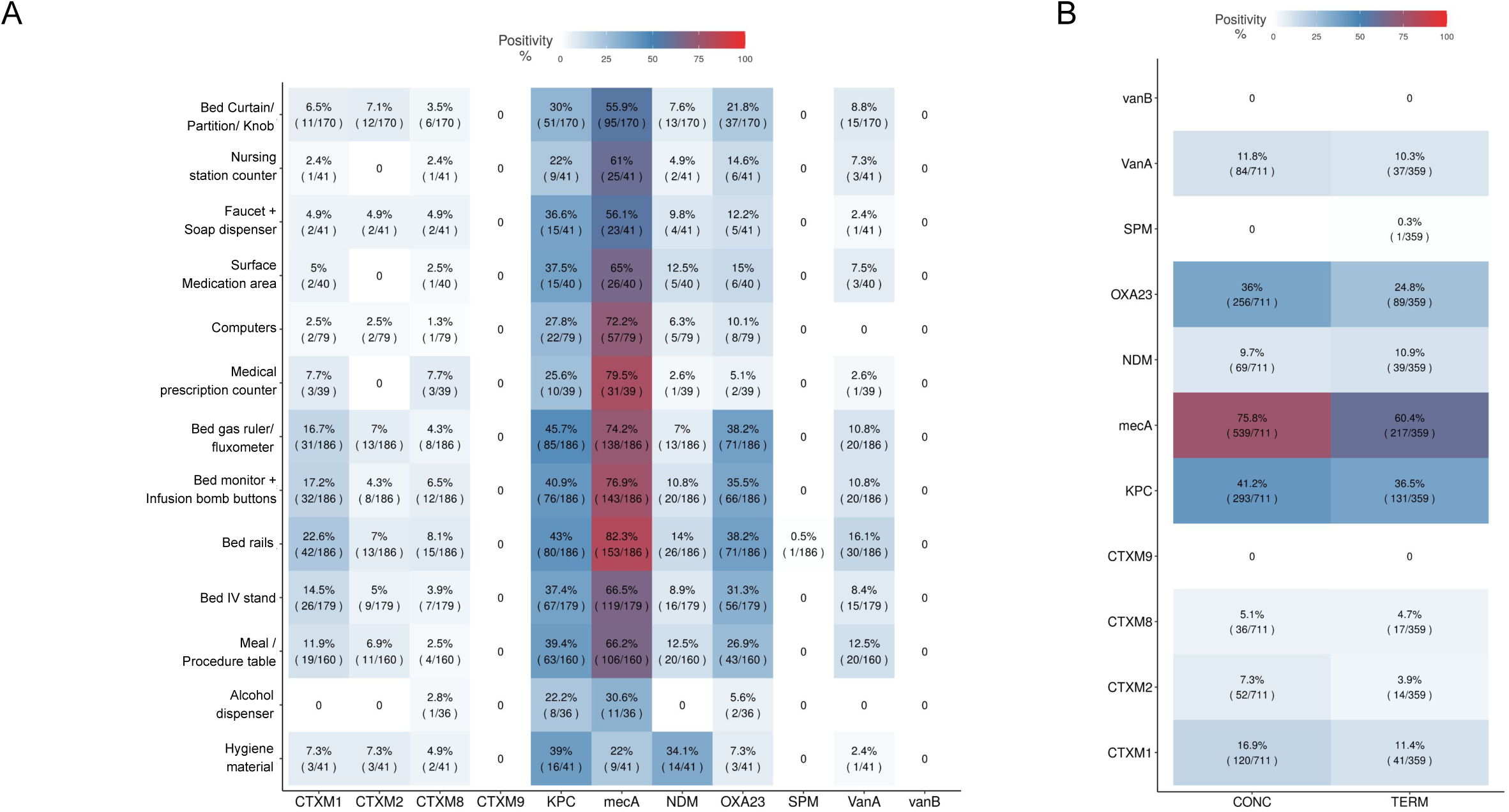
Antimicrobial resistant genes positivity among surface samples and cleaning processes. (A) Hospital sampled surfaces and AMR genes positivity proportions considering the total amount of samples collected for each surface type. (B) Resistance genes positivity in ICU bed samples considering the kind of cleaning process used: CONC (daily concurrent cleaning while patient is still in bed) and TERM (terminal cleaning after patient discharge). Positivity rates are also indicated by color scales from clear blue to red (0 to 100%).

In a comparative analysis, hospital samples were grouped by their Brazilian states of origin and the prevalence AMR rate was evaluated by country state (**Fig. 7A**). *mecA* and *bla*_KPC-like_ AMR genes were the most prevalent ones, reaching more than 70% prevalence values. Most prevalent genes: *bla*_CTX-M-1_ group, *bla*_CTX-M-2_ group, *bla*_CTX-M-8_ group, *bla*_NDM-like_, *bla*_OXA-23-like_, *vanA*, *bla*_KPC-like_ and *mecA* were represented in Brazilian maps by their percentage of positivity in that state collected samples (**Fig. 7B**). However, one must be careful, since these results may not demonstrate the complete reality of the Brazilian states, it is necessary to consider that hospital sampling was not equally distributed among states, some are well more represented than others. Thus, this data is only related to the profile of collected samples in this study. AMR genes positivity rates from individual hospitals can be found in **Supp.fig. 6**.

**Figure 7.**
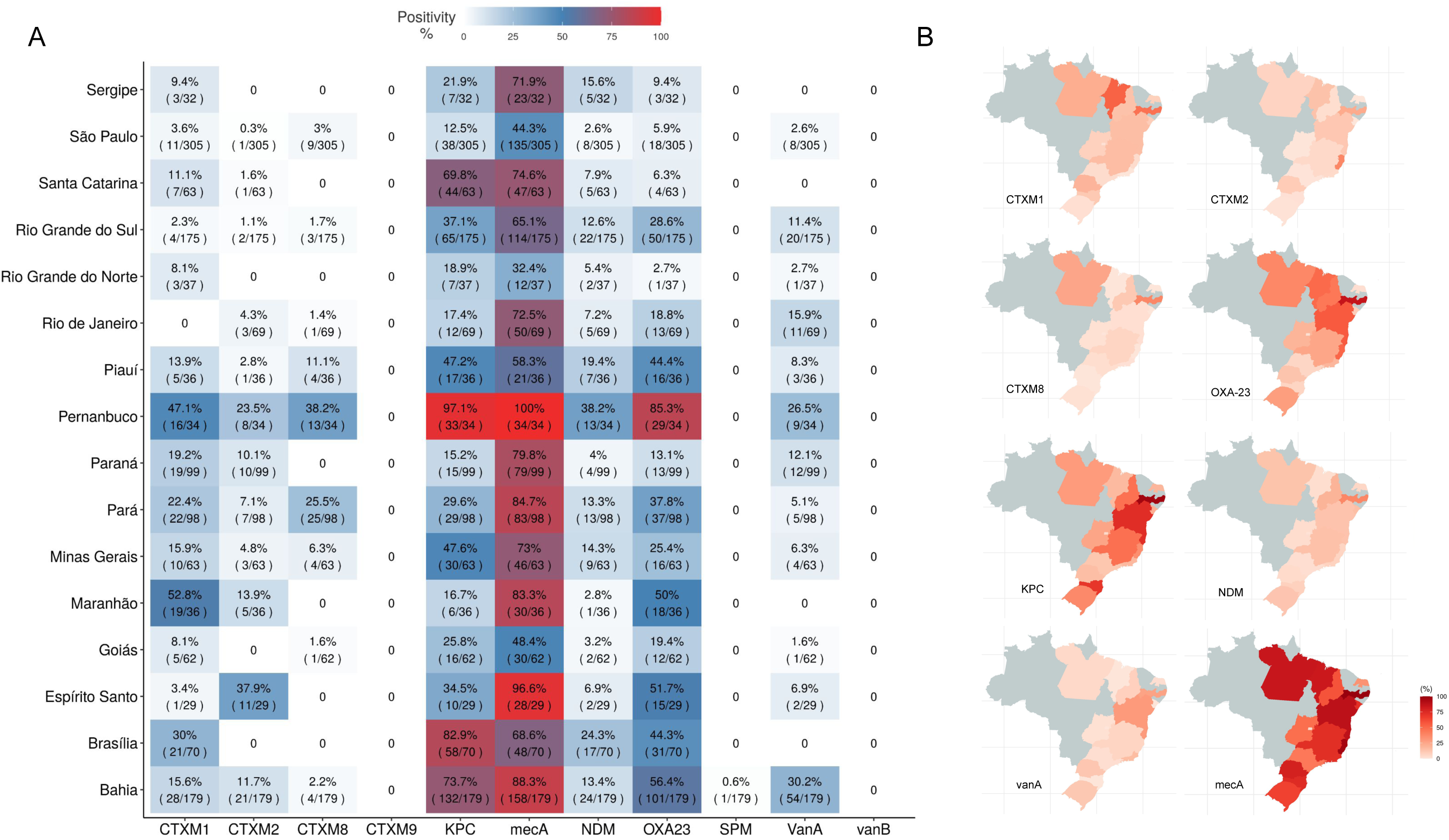
Antimicrobial resistant genes distribution around Brazilian states. AMR detected genes were grouped by hospital location in the country and their proportions of positivity is shown in a heatmap (A) and also individually highlighted by state in country maps for the most abundant genes, *bla*_CTX-M-1_ group, *bla*_CTX-M-2_ group, *bla*_CTX-M-8_, *bla*_OXA-23-like_, *bla*_KPC-like_, *bla*_NDM-like_, *vanA* and *mecA* (B).

## Discussion

A comprehensive analysis of 41 hospitals and its sanitizers used by the hygiene and nursing teams in ICUs over 16 states around Brazil was carried out to identify the bacterial profiling of hospital environments and its AMR genes. In summary, a very heterogeneous scenario was found across the country in terms of microbiome. Clusters of hospitals could be found, but they did not follow any geographic pattern (e.g., Brazilian states). Diversity patterns by region seem to be present, although not so relevant and difficult to interpret. Contamination of sanitizers were not infrequent, correspondence with the environment was found, but the clinical relevance of that is still to be defined.

The high-throughput amplicon sequencing used in this study enabled a detailed analysis of bacterial taxa in the hospital surfaces, as already performed in several other studies (18,19,23,35). Our results showed a wide diversity of microbial populations among most hospitals, with certain bacteria being present in all of them, including ones related to healthcare-associated infections. Some of the most represented microorganisms are also detected by other studies in Brazilian ICU surfaces, such as *Staphylococcus spp, Pseudomonas spp, Acinetobacter spp* and *Bacillus spp* (16,17,36). Considering the total bacterial load in surfaces, the majority could be classified as hospital pathogens. Previous studies have demonstrated that total sequence reads from high-throughput amplicon sequencing, using an equivolumetric library preparation methodology, allow bacterial load estimation in the collected samples (26,37). This finding emphasizes the importance of hospital microbiome profiling as a tool in understanding and controlling BMR horizontal transmission.

Hierarchical clustering of bacterial profiles grouped different hospitals and ICU sample sites according to their major bacterial positivity rates. These groupings among hospitals were also reinforced in the beta-diversity bacterial analysis. São Paulo’s hospitals have the most widely distributed bacterial profile in beta-diversity results, maybe related to the high level of flow of people from all over the country in this state. It was demonstrated that, in fact, flow of people can modify the hospital and ICU environmental microbiome (18,38). However, despite the observed clustering patterns, no correlation was found between the bacterial grouping of hospitals and the metadata analyzed, such as hospital geographical locations, or the sanitization products used. A deeper survey of hospital metrics and indicators must be performed to understand the similarity patterns, or if they are just random.

Inanimate surfaces and equipment in ICU have currently been shown as bacterial contaminated sites that may contribute to patients acquired colonization or infection (39,40). While every hospital in our study exhibits a distinct bacterial profile, there seems to be a recurring trend related to sampling sites. Hygiene materials, soap dispenser and faucet are the least contaminated samples, while bed sites as gas ruler, monitor, infusion bomb and bed rails were more contaminated, with higher bacterial positivity rates. Common use areas as medical prescription and nursing station counter, as well as common use tables (procedure and meal table) were more related considering some hospital pathogens positivity rates. The contamination of frequently touched hospital surfaces with drug-resistant bacteria such as methicillin-resistant *Staphylococcus aureus* (MRSA), vancomycin-resistant *Enterococcus* (VRE), carbapenem-resistant Enterobacteriaceae (CRE), and others microorganisms has been well documented (41). Undoubtedly, high-touch surfaces, such as areas near the patient or frequently touched by healthcare workers, may represent ‘critical surfaces’ due to their potential for cross-transmission of pathogens, and these surfaces may also benefit from routine cleaning with disinfectants (42).

The prevalence of antimicrobial resistance genes in hospital surface samples was notable, with the *mecA*, *bla*_KPC-like_, *bla*_NDM-like_, and *bla*_OXA-23-like_ genes being the most frequently detected. Those results are congruent when looking at the microbial profile found in the hospitals, with a high prevalence of nosocomial pathogens, that may carry these resistant genes. Also, *bla*_NDM-like_ has been associated with multidrug resistance and has been reported from various Brazilian regions in different gram-negative species (43,44). *bla*_KPC-like_ has also been associated with MDR (45), and it has also been found in other taxa apart from *Klebsiella pneumoniae*, such as *Acinetobacter baumannii* and *Pseudomonas spp* (46,47), highlighting the importance of comprehensive surveillance and control measures at regional level. As stated before, hospitals with higher bacterial load are the ones with a higher presence of nosocomial pathoges, and, as consequence, the ones with more positive samples for AMR genes. However, even though our results show a high level of AMR genes among hospitals in Brazil, it is also important to consider that the samples for this study were collected between 2020-2021, when SARS-CoV-2 pandemic was widely spread around the world, and the hospitals were overcrowded, and the health professionals were overloaded. Previous studies have shown that hospitals were suffering with a higher level of HAIs during this period (48,49). Furthermore, hospital sampling was not equally distributed among all Brazilian regions, some regions are well more represented than others.

The *mecA* gene is a crucial biomarker of methicillin resistance and holds significant importance in the context of antimicrobial resistance and healthcare, and it was one with the highest detection rate in most analyzed hospitals. Previous studies have shown that the presence of the *mecA* gene is not limited solely to *Staphylococcus aureus*; it has also been identified in other species within the same genera, including *Staphylococcus epidermidis* (50,51), which has already been previously reported in Brazilian hospital (17). *S. epidermidis* was present in all the hospitals analyzed in this study, usually in a high prevalence among samples. It is highly possible that the high level of *mecA* gene detected in this study is related to *S. epidermidis* prevalence, as *S. aureus* is not among the most detected taxa, nor was detected by DNA high-throughput sequencing in all hospitals. In addition, as *S. epidermidis* may also be a contaminant (and not always pathogen), its relevance is difficult to be accessed.

The investigation on in-use hospital sanitizers revealed a wide variety of products and active principles being used across different hospitals. The aim of this investigation was not to test the sanitizer efficacy itself, but rather evaluate its effectiveness in routine use by the hygiene and nursing teams (process) and identify patterns in sanitizer used around the different regions of the country. Indeed, there is no pattern in sanitizer used around the analyzed hospitals, as 42 different products were being used by the time this study was made, and, among them, 18 different active principles. In this study, only three hospitals were using the same active principle sanitizer by both nursing and hygiene team, and the bacterial profile among these hospitals was not similar, indicating that the sanitizer active principle was not a definitive limitant for bacterial diversity. Furthermore, regardless of the active principle, it is not possible to find correlations between them and bacterial profiling, HAI-profiling or AMR genes detection; each hospital has its own bacteriome, despite the sanitizer used, at least in this first survey. The variation in products suggests a lack of clarity about what types of products are most efficient in reducing the risk of infection for patients and using different or several products and assigning varying responsibilities for cleaning within a hospital can lead to confusion and inappropriate use of disinfectants, including under use, overuse and interaction of products that are not designed to be used concurrently (42). Another study in a Brazilian hospital also evaluated the bacterial profile after the cleaning process and detected several HAI-genera following sanitization in the ICU (36).

One significant observation in this study was that viable bacterial cells were indeed detected in some in-use sanitizers, so it is important to ensure that the teams are using proven efficacy sanitizers, in correct dilutions and procedures, that are able to kill bacterial cells, as a decrease in HAIs through improved cleaning practices and the use of corresponding disinfection methods can be reached (52–54). Environmental cleaning products (detergents and disinfectants) are often sold as concentrated formulas that are diluted (i.e. combined with water) to create a solution. This process must be strictly controlled and professionals must be trained and preferably use automated dosers for dilution. Furthermore, they must comply with the expiration date of the solutions after dilution and store them in clean, closed containers (55,56). The use of detergents (i.e., soap and water) versus disinfectant chemicals has been an area of controversy. Detergent solutions have the potential to become contaminated with bacteria during the cleaning process, which can result in further spread of bacteria across surfaces and diluted products have a greater risk of inadequacy if the rules are not followed (57). Despite product contamination, as demonstrated by *Serratia marcescens* and *Achromobacter xylosoxidans* presence in a quaternary ammonium disinfectant and its cleaned surfaces (58), the cleaning process should also be effective and standardized. Other studies demonstrated carryover contamination by cleaning wipes when the process is not well established (59) and when there is a greater compliance in the cleaning process by healthcare workers, it was possible to drastically reduce HAI caused by *C difficile,* MRSA, and VRE (60). The presence of nosocomial pathogens in some sanitizer samples raises concerns about their effectiveness in controlling bacterial growth, and the detection of antimicrobial resistance genes in sanitizer samples further emphasizes the importance of assessing the efficacy of sanitization protocols.

## Conclusion

This study has assessed the bacterial profile and AMR genes of upper middle-income country hospitals in different regions, demonstrating a variety and the spreading of healthcare-associated infection bacteria and antimicrobial resistance genes around the country. The importance of understanding bacterial profiles, and hospital clusters regarding ICU/hospital environment microbiome for implementing targeted interventions to control HAIs and antimicrobial resistance, and the meaning and impacts of sanitizer contamination in terms of HAI dissemination should be addressed in future studies.

## Data Availability

All sequence data are deposited in NCBI BioProject SUB13793553

## Author Contributions

D.C.B processed samples, analyzed the results and wrote the manuscript; C.V.S. was responsible for data collection and involved in the study design; A.P.C was involved in the study design, sample processing, data analysis and manuscript writing; G.N.F.C. performed bioinformatic analysis; L.D.T. and L.S.R.A contributed to the discussion of the results; B.T, A.B.C., B.A and F.T.P. were responsible for the grant and for the IMPACTO MR platform, contributed to the study design and for the discussion; L.F.V.O. contributed to the study design and data analysis; A.J.P. was responsible for the grant and for the IMPACTO MR platform, planned and designed the study, and contributed directly to the manuscript writing. All the authors revised and approved this final version of the text.

## Acknowledgements

We are indebted with Secretaria de Ciência, Tecnologia e Insumos Estratégicos/SCTIE (Brazilian Ministry of Health) and the Coordenação de Saneantes (COSAN) da Gerência de Produtos de Higiene, Perfumes, Cosméticos e Saneantes (GHCOS) da Agência Nacional de Vigilância Sanitária / ANVISA (Brazilian Health Regulatory Agency) for the technical and administrative support. Institutional Development Support Program (Programa de Apoio ao Desenvolvimento Institucional do Sistema Único de Saúde / PROADI-SUS). We thank Prof. Dr. Ana Cristina Gales (ALERTA Laboratory, Universidade Federal de São Paulo, USP) who kindly provided positive control strains for the resistance gene (RGene) study.

## Supplementary Material

**Supplementary figure 1.** Bacterial positivity rates for each hospital’s most abundant bacteria in different surface collection sites.

**Supplementary figure 2.** Bacterial positivity rates for a group of 17 HAI-related bacteria in different surface collection sites for each hospital.

**Supplementary figure 3.** Bacterial alpha-diversity median values for Shannon (A) and Richness (B) indexes considering each hospital analyzed.

**Supplementary figure 4.** General beta-diversity profiles and decomposed by hospital identification.

**Supplementary figure 5.** General beta-diversity profiles and decomposed by Brazilian states origin of samples.

**Supplementary figure 6.** Antimicrobial resistance genes analyzed and positivity rates by hospital.

**Supplementary table 1.** Hospital collection sites. *Samples were collected 5 times (from 5 different beds, 3 in rooms with patients, and 2 vacant rooms, after terminal cleaning).

**Supplementary table 2.** List of hospital sanitizers.

**Supplementary table 3.** Microbiology culturing, 16S rRNA amplicon sequencing from media grown bacteria, and taxonomical identification, and AMR genes analysis results for hospital sanitizers with real-time PCR (RGene).

## Data Availability Statement

All sequence data are deposited in NCBI BioProject SUB13793553

