## Supplementary figures and images for "A transversal overview of Intensive Care Units environmental microbiome and antimicrobial resistance profile in Brazil"

### Fig S1

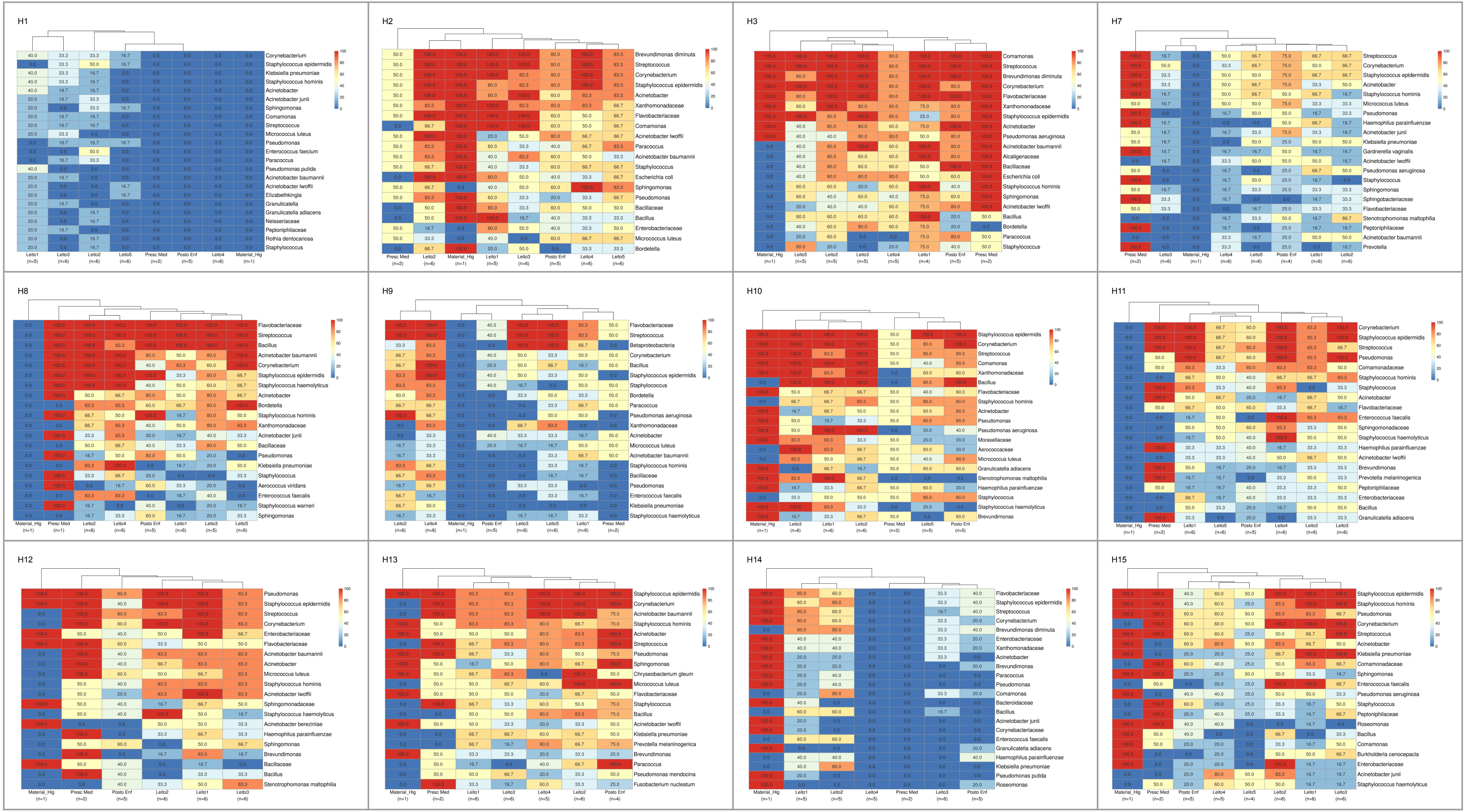

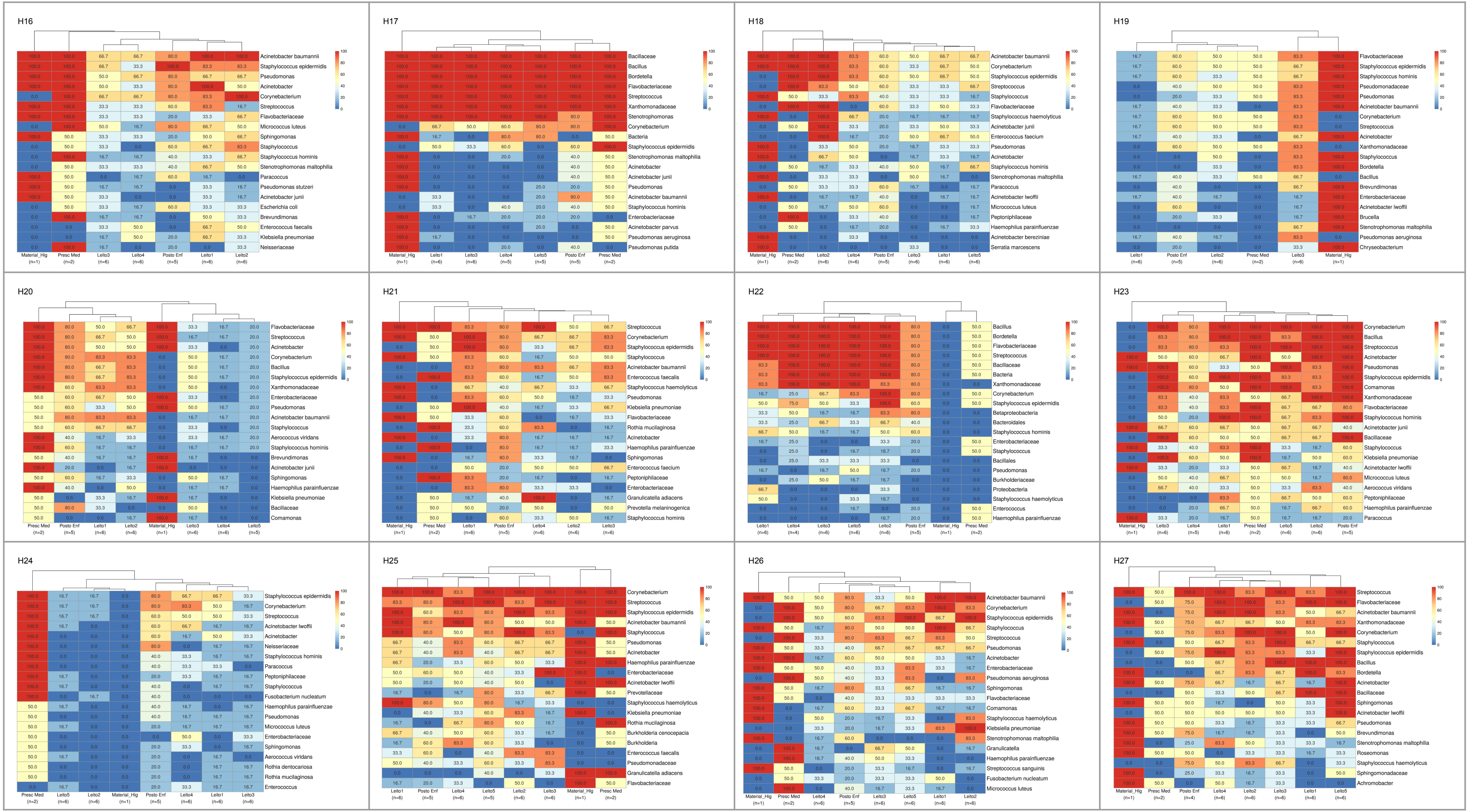

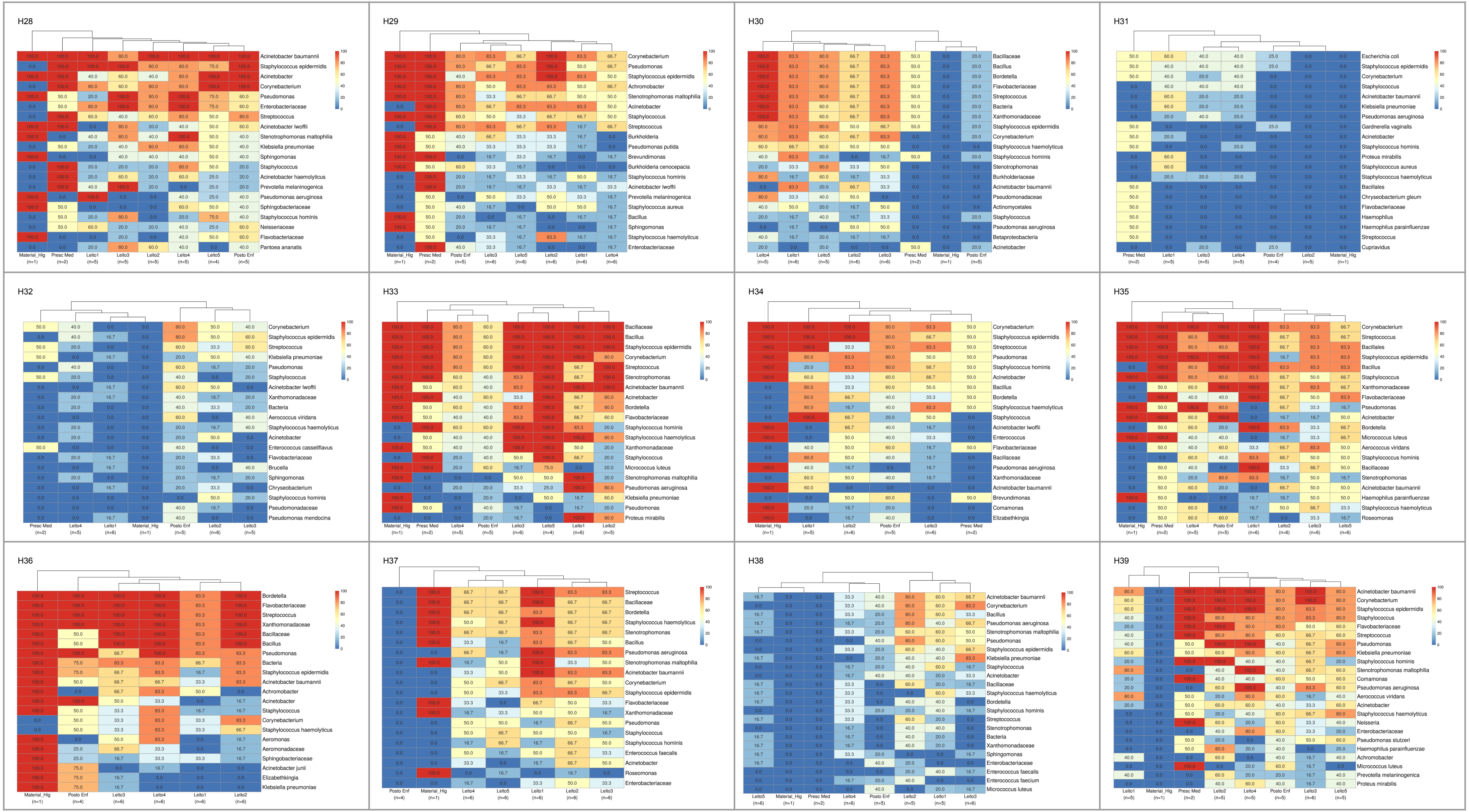

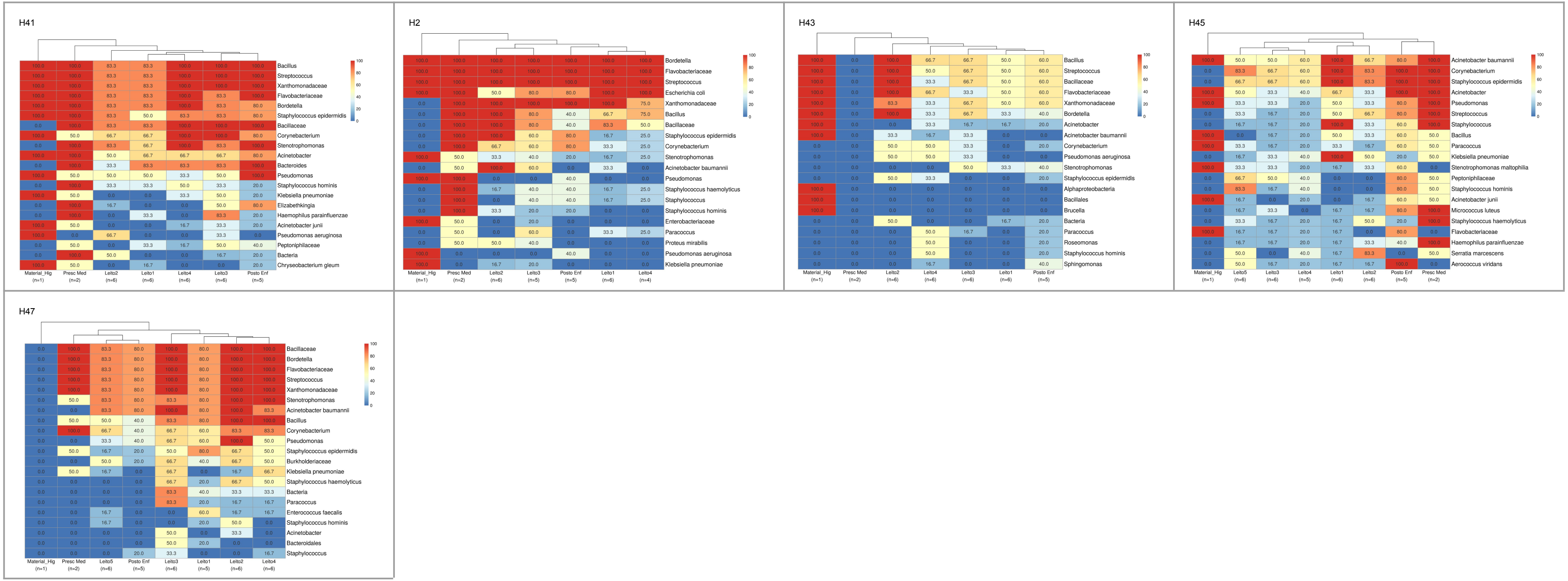

### Fig S2

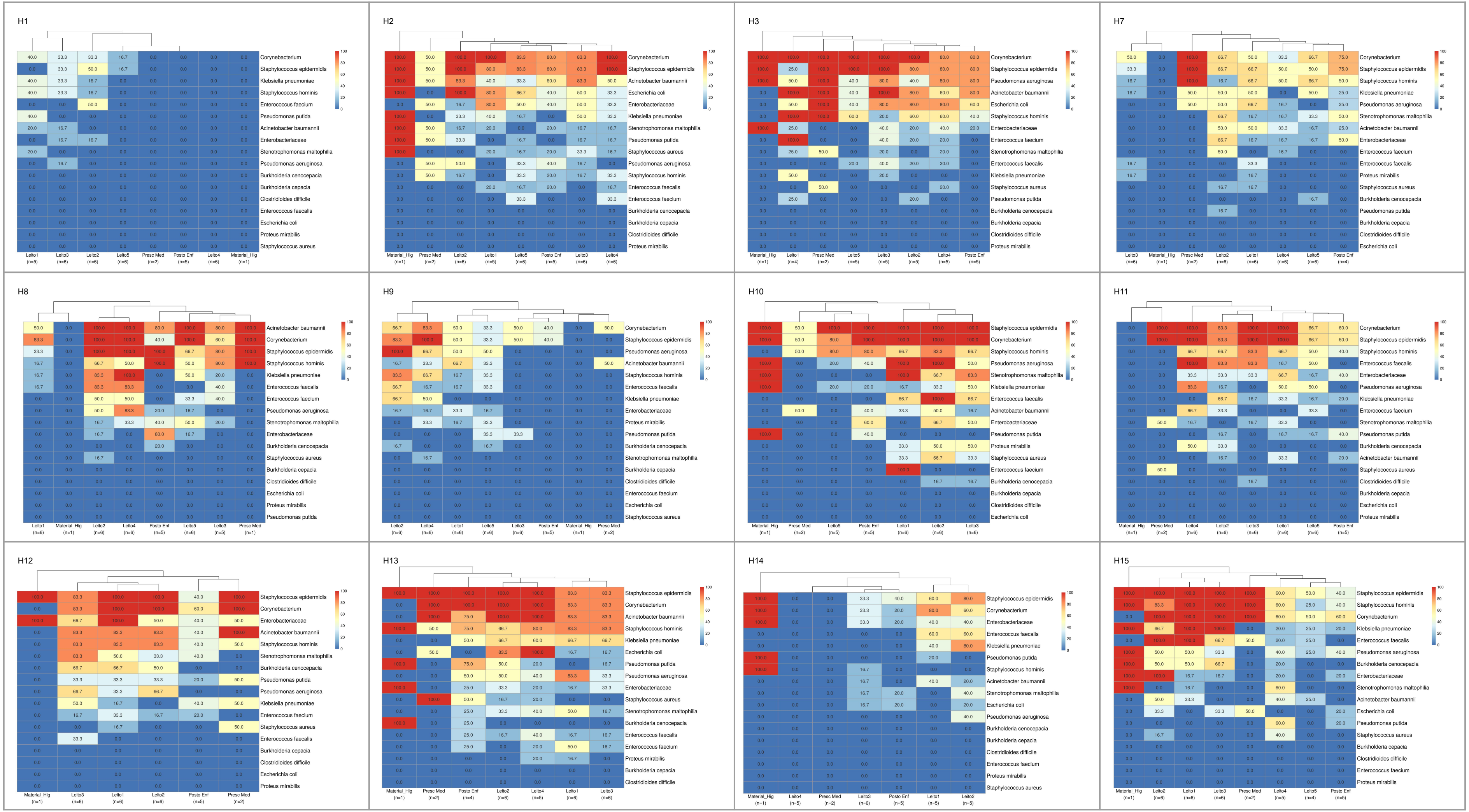

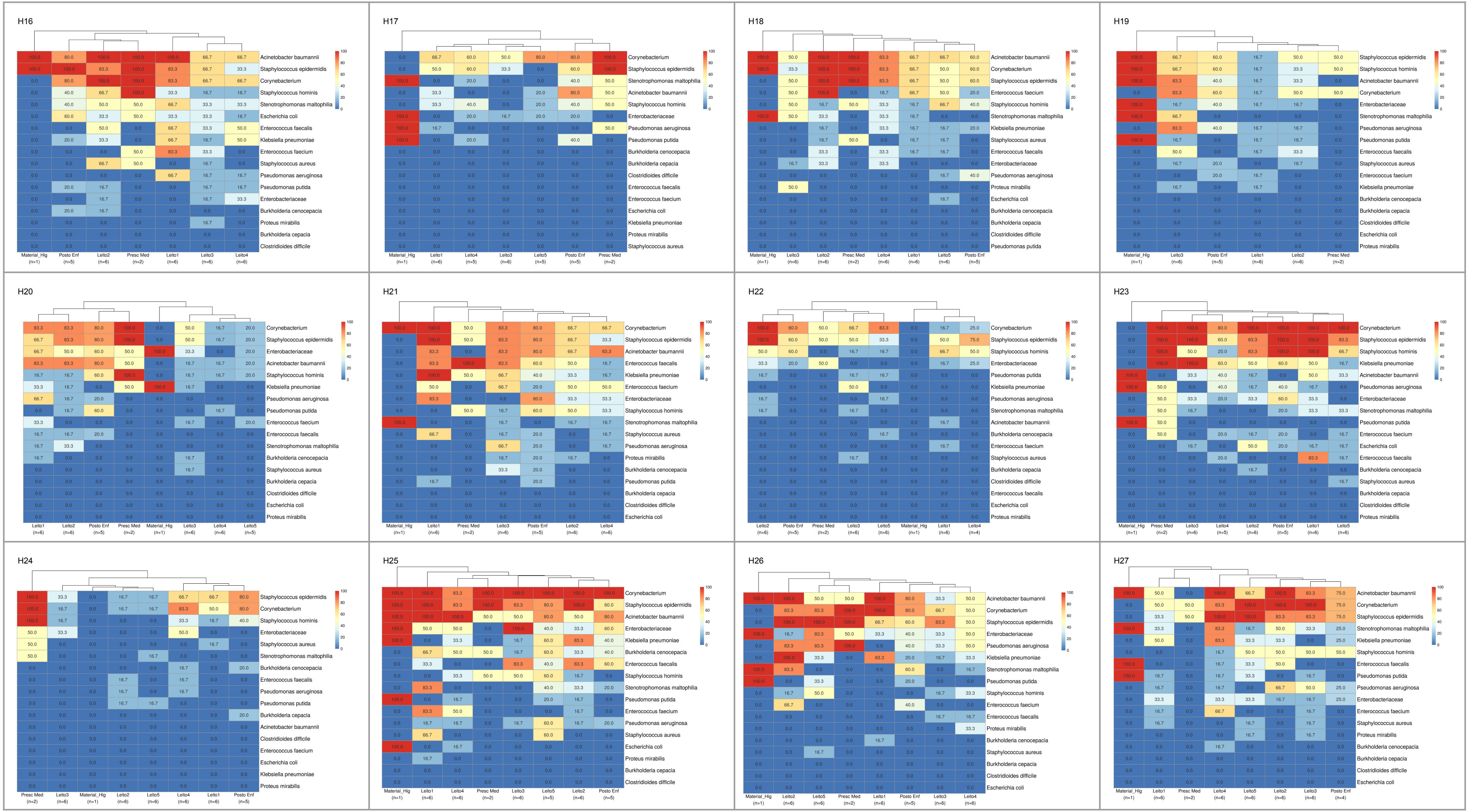

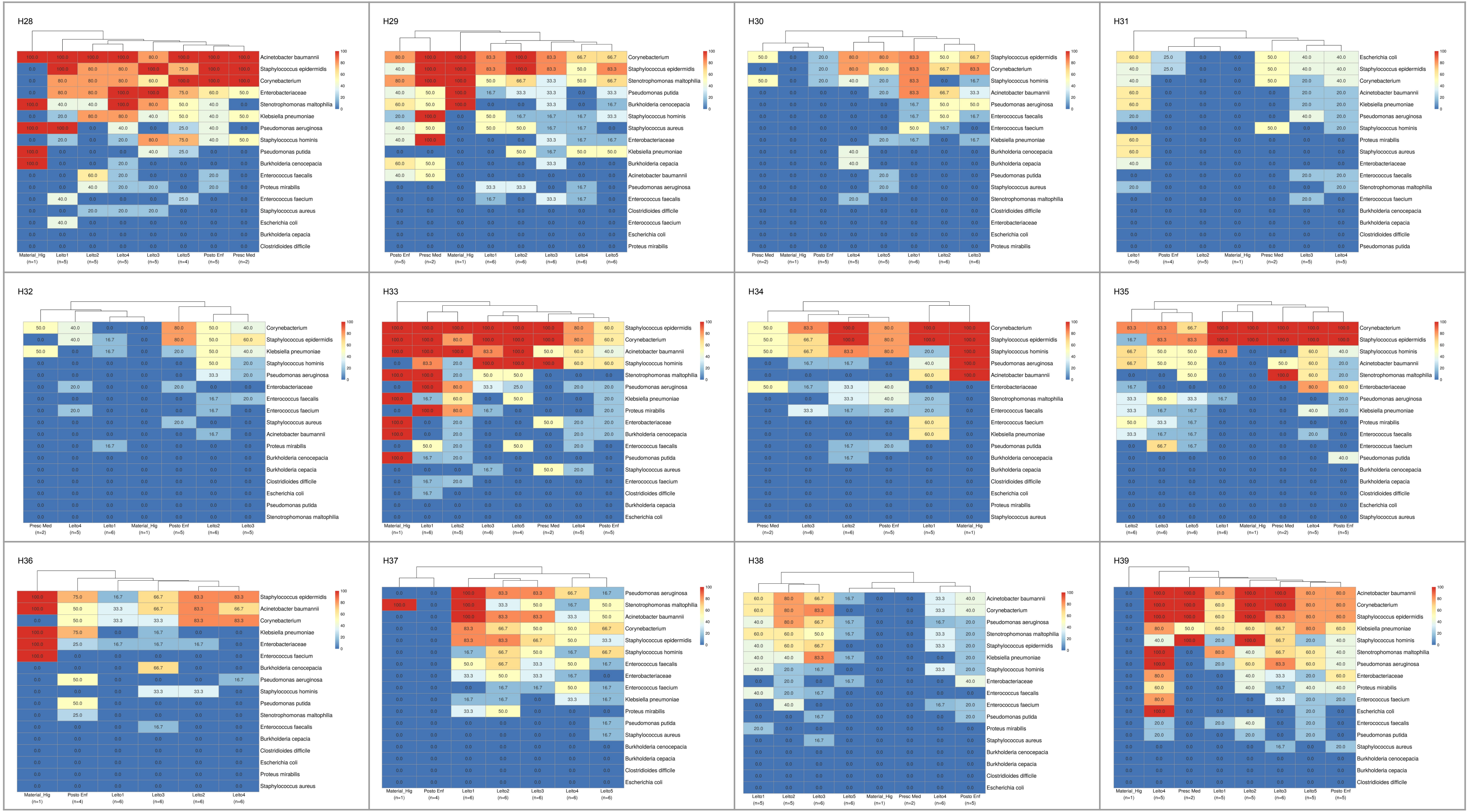

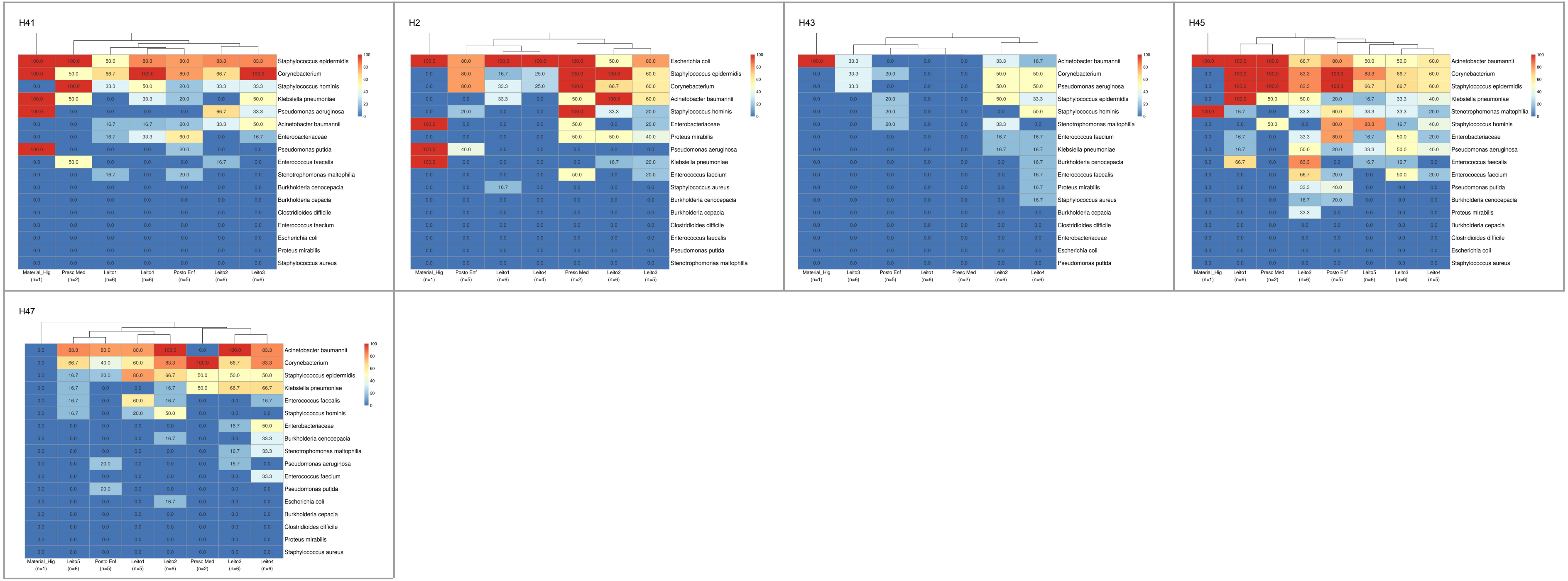

### Fig S3

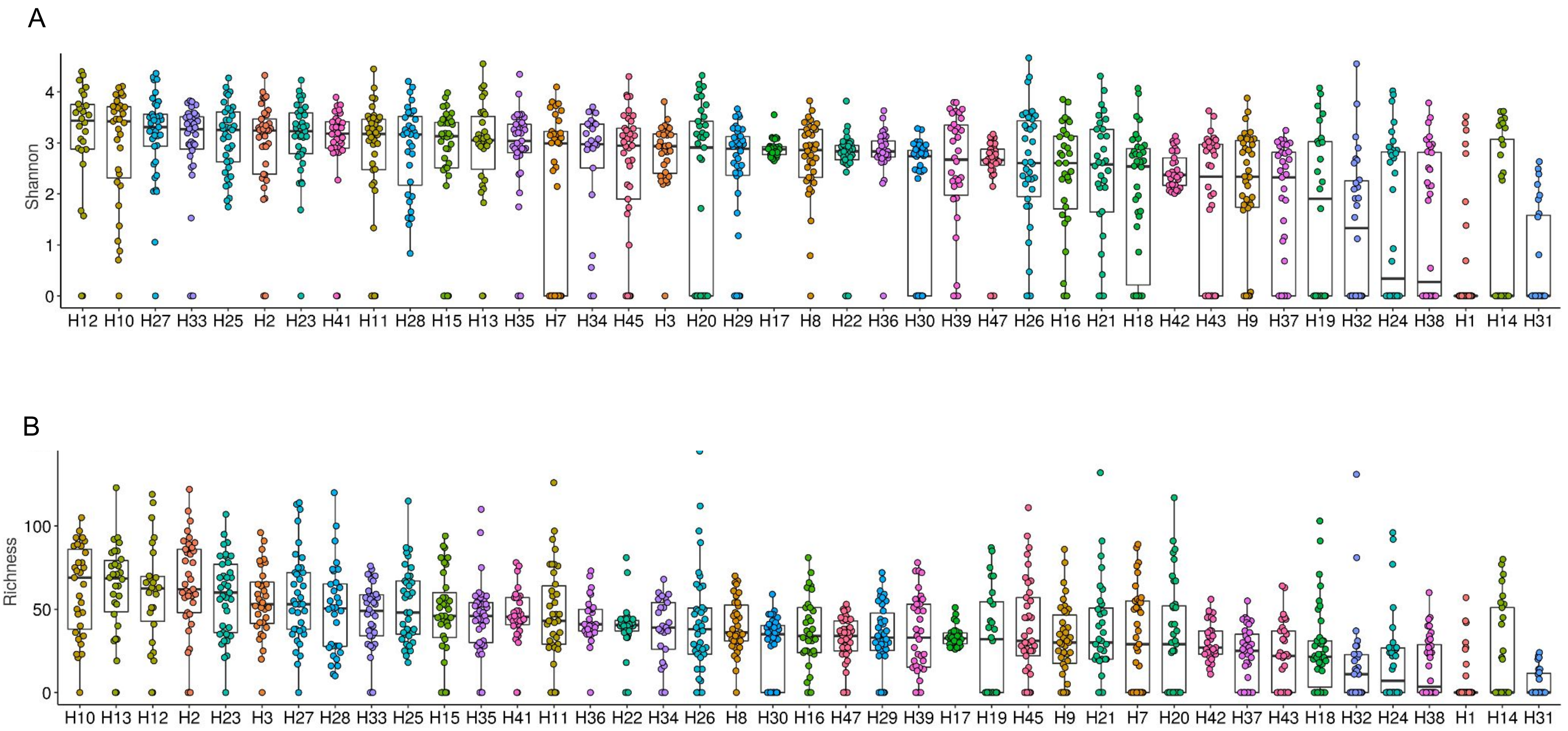

### Fig S4

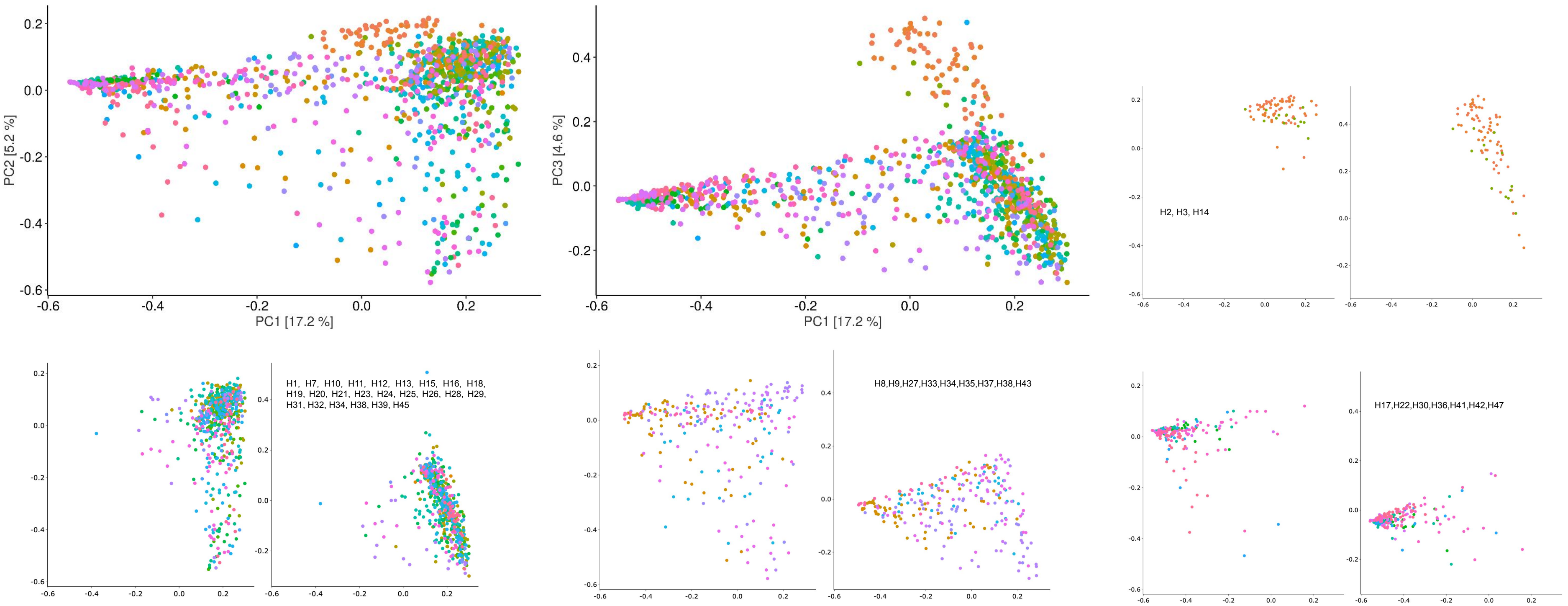

### Fig S5

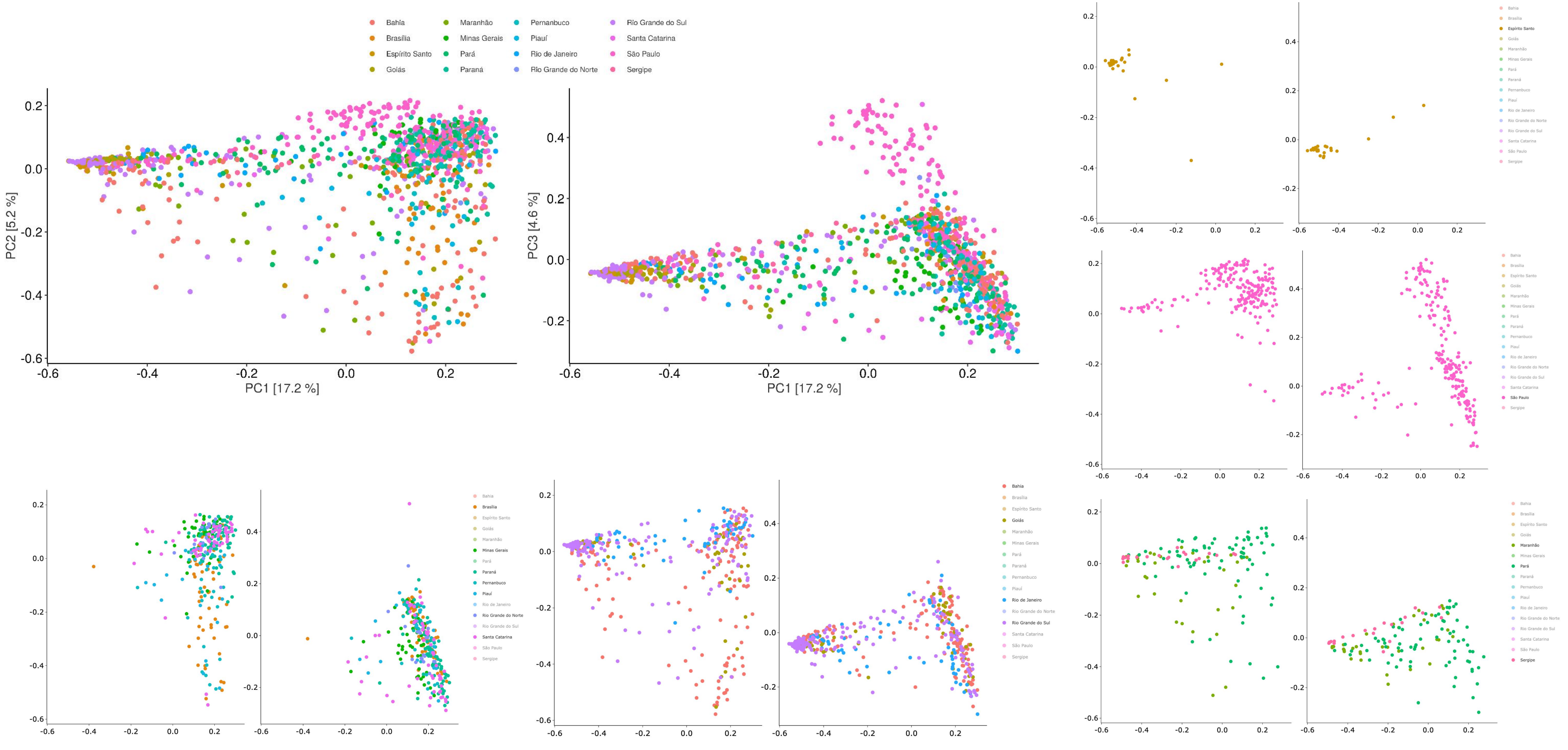

### Fig S6

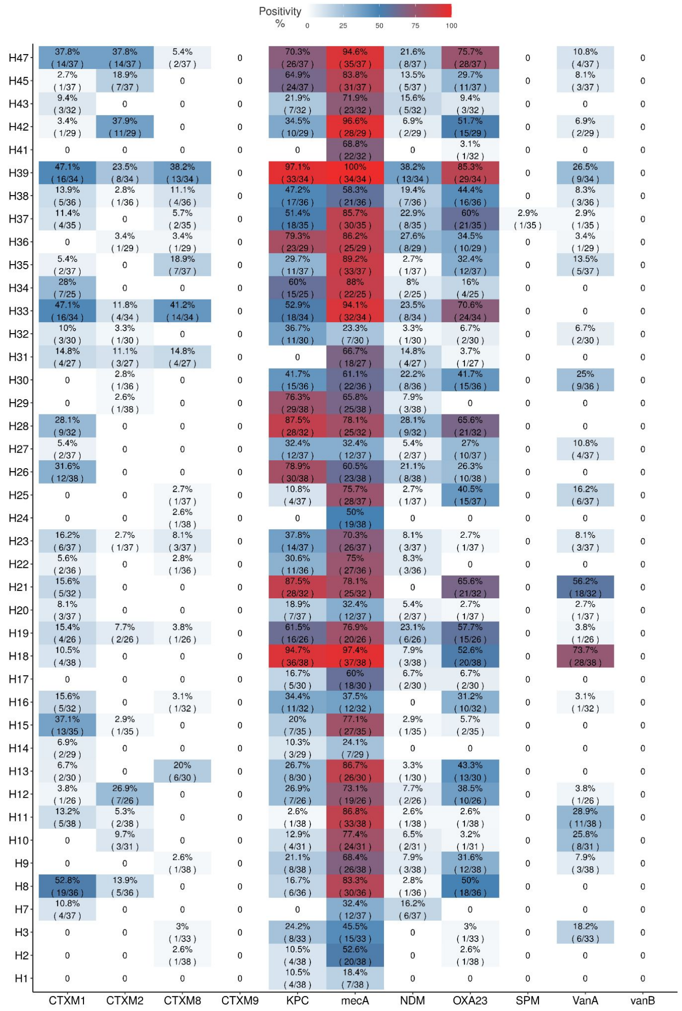
